# Autoantibodies to Arginine-rich Sequences Mimicking Epstein-Barr Virus in Post-COVID and Myalgic Encephalomyelitis/Chronic Fatigue Syndrome

**DOI:** 10.1101/2024.12.30.24319800

**Authors:** Friederike Hoheisel, Kathrin Maria Fleischer, Kerstin Rubarth, Nuno Sepúlveda, Sandra Bauer, Frank Konietschke, Claudia Kedor, Annika Elisa Stein, Kirsten Wittke, Martina Seifert, Judith Bellmann-Strobl, Josef Mautner, Uta Behrends, Carmen Scheibenbogen, Franziska Sotzny

**Affiliations:** Institute of Medical Immunology, Charité – Universitätsmedizin Berlin, Corporate Member of Freie Universität Berlin and Humboldt Universität zu Berlin and Berlin Institute of Health, Berlin, Germany; Institute of Virology, Helmholtz Munich, Germany; Children’s Hospital, School of Medicine, Technical University of Munich, Munich, Germany; Institute of Biometry and Clinical Epidemiology, Charité – Universitätsmedizin Berlin, Corporate Member of Freie Universität Berlin, Humboldt-Universität zu Berlin, Berlin, Germany; Faculty of Mathematics & Information Science, Warsaw University of Technology, Varsóvia, Polónia; CEAUL - Centro de Estatística e Aplicações da Universidade de Lisboa, Lisboa, Portugal; Berlin Institute of Health at Charité – Universitätsmedizin Berlin, BIH Center for Regenerative Therapies (BCRT), Berlin, Germany; DZHK (German Center for Cardiovascular Research), partner site, Germany; Experimental and Research Center (ECRC), Charité - Universitätsmedizin Berlin, Corporate Member of Freie Universität Berlin, Humboldt Universität zu Berlin and Berlin Institute of Health, Berlin, Germany; NeuroCure Research Centre, Charité - Universitätsmedizin Berlin, Corporate Member of Freie Universität Berlin, Humboldt Universität zu Berlin and Berlin Institute of Health, Berlin, Germany; German Center for Infection Research (DZIF), Berlin, Germany; Technical University of Munich, Munich, Germany

**Keywords:** autoantibodies, cross-reactivity, EBV, arginine-rich peptides, post-COVID syndrome, ME/CFS

## Abstract

**Background:** Epstein-Barr virus (EBV) infection is a known trigger and risk factor for myalgic encephalomyelitis/chronic fatigue syndrome (ME/CFS) and post-COVID syndrome (PCS). In previous studies, we found enhanced IgG reactivity to EBV EBNA4 and EBNA6 arginine-rich sequences in postinfectious ME/CFS (piME/CFS).

**Objective:** This study aims to investigate IgG responses to arginine-rich (poly-R) EBNA4 and EBNA6 sequences and homologous human sequences in PCS and ME/CFS.

**Methods:** The IgG responses against poly-R EBNA4 and EBNA6 and corresponding homologous human 15-mer peptides and respective full-length proteins were analyzed using a cytometric bead array (CBA) and a multiplex dot-blot assay. Sera of 45 PCS patients diagnosed according to WHO criteria, with 26 patients fulfilling the Canadian Consensus criteria for ME/CFS (pcME/CFS), 36 patients with non-COVID post-infectious ME/CFS (piME/CFS), and 34 healthy controls (HC) were investigated.

**Results:** Autoantibodies to poly-R peptide sequences of the neuronal antigen SRRM3, the ion channel SLC24A3, TGF-β signaling regulator TSPLY2, angiogenic regulator TSPYL5, as well as to full-length α-adrenergic receptor (ADRA) proteins were more frequent in patients. Several autoantibodies were positively associated with key symptoms of autonomic dysfunction, fatigue, cognition, and pain.

**Conclusion:** Collectively, we identified autoantibodies with new antigen specificities with a potential role in PCS and ME/CFS.

**Clinical Implication:** These finding should prompt further studies on the function of these autoantibodies, their exploitation for diagnostic use, and of drugs targeting autoantibodies.

**Capsule summary:** Our study reveals elevated autoantibodies to EBV-related poly-R sequences and their human homologues in PCS and ME/CFS patients associated with symptom severity, suggesting a potential role in disease pathogenesis.

## 1 Introduction

Severe Acute Respiratory Syndrome Coronavirus 2 (SARS-CoV-2) infection has been shown to induce a range of autoantibodies (1–3). Several studies have demonstrated antinuclear autoantibodies (ANA), as well as autoantibodies directed against G protein-coupled receptors (GPCRs), and various neuronal, muscular, and other intra-and extracellular proteins in post-COVID syndrome (PCS). There is evidence of sequence similarities to SARS-CoV-2 proteins (3). Autoantibodies have also been found to be associated with key symptoms of the disease (1, 3–7). Importantly, two recent studies have shown that transferring IgG from PCS patients can induce similar symptoms in mice (8, 9). In the study by Santos Guedes de Sa *et al*., patients showed a broad pattern of autoantibodies reactive to neuronal tissues and meninges. IgG from individual patients induced distinct symptoms such as pain, hypersensitivity or loss of coordination in mice.

A subgroup of PCS patients fulfill the diagnostic criteria of myalgic encephalomyelitis/chronic fatigue syndrome (ME/CFS), with the key symptom of exertional intolerance and worsening of symptoms after daily activity, known as post-exertional malaise (PEM) (10, 11). There is also increasing evidence for the role of autoantibodies in post-infectious ME/CFS (12, 13). GPCR autoantibodies have been found to be elevated in a subgroup of ME/CFS and to correlate with symptom severity and alterations in magnetic resonance imaging (MRI) indices (14–17). Epstein-Barr virus (EBV) is a well-known trigger for ME/CFS, and EBV reactivation during COVID-19 is a risk factor for PCS (18–20). Altered antibody responses to EBV antigens have been identified in ME/CFS, and there is first evidence of cross-reactivity with human proteins. A recent study identified autoantibodies to myelin basic protein in ME/CFS, a known autoantigen in multiple sclerosis with sequence similarity to EBV Epstein-Barr nuclear antigen (EBNA) 1 (21). In a previous study, we analyzed IgG responses against more than 3000 overlapping 15-mer peptide sequences derived from 14 EBV proteins. We observed elevated IgG responses against various peptides in ME/CFS patients (22). Among these, we found enhanced IgG reactivity against several peptides in an EBNA6 repeat sequence with homologies to several human proteins and an increased IgG reactivity against the full-length EBNA6 protein. An extended bioinformatic data analysis revealed an enhanced IgG reactivity against an arginine-rich (poly-R) sequence in the EBNA6 sequence and in EBNA4 among piME/CFS patients. These sequences showed homologies with several human proteins (**23, 24**).

In the present study, we analyzed IgG reactivity to peptides of the poly-R EBV EBNA4 and 6 and homologous human sequences, as well as corresponding proteins in PCS and ME/CFS. We further examined the association between titers of these antibodies and key clinical symptoms.

## 2 Methods

### 2.1 Study participants

For this exploratory study, female participants aged 18 to 59 years were recruited at the Charité Fatigue Centre, Berlin. Serum samples were collected from 45 PCS patients suffering from moderate to severe fatigue with 26 of them fulfilling the Canadian Consensus Criteria (CCC) for the diagnosis of ME/CFS (pcME/CFS). In the case of 2 of 26 pcME/CFS patients and 3 of 19 PCS patients with a disease duration of less than 6 months, the diagnosis was confirmed at month 6. In addition, we included serum samples from 36 other postinfectious ME/CFS (piME/CFS) patients and from 34 age-matched female HCs.

Detailed cohort information is displayed in Table I. Disease, and symptom severity were assessed in appropriate questionnaires. The functional disability was assessed with the Bell score, ranging from 0 to 100 (with 100 for no restrictions) (25). Physical function and daily activities were evaluated using the Short Form Health Survey 36 (SF-36) ranging from 0 to 100 (greatest to no restrictions) (26). PEM severity was assessed according to the brief DSQ-PEM questionnaire and scores ranging from 0 to 46 calculated (no to frequent/severe PEM) (27). The severity of the key symptoms, fatigue, pain, and cognitive impairment was quantified using a Likert scale (1 = no symptoms to 10 = severe symptoms). Autonomic dysfunction was assessed using the Composite Autonomic Symptom Score 31 (COMPASS-31), ranging from 0 to 100 (no to strongest impairment) (28).

**Table I.**
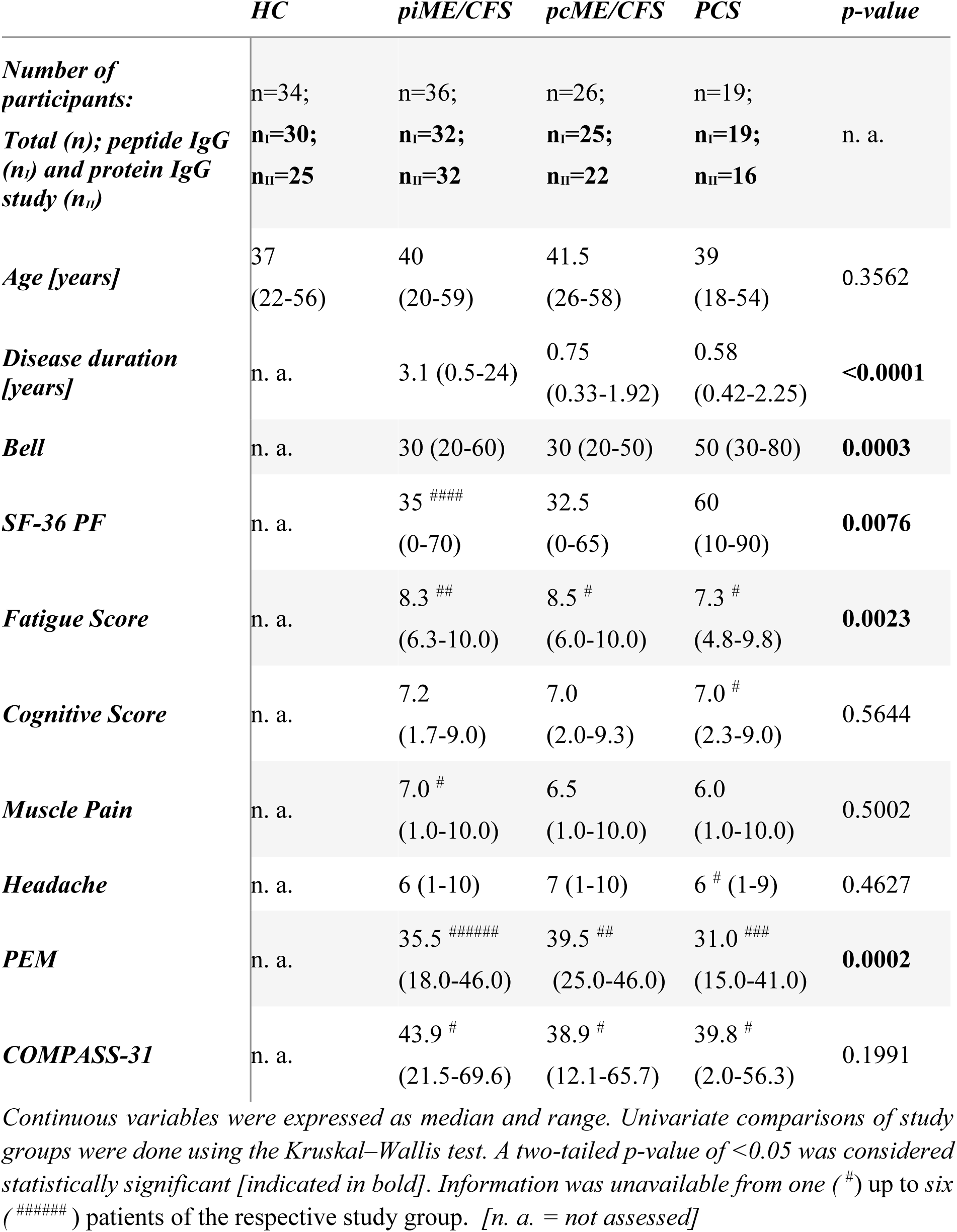
Cohort characteristics.

Routine laboratory parameters were determined at the Charité diagnostics laboratory Labor Berlin GmbH (Berlin, Germany). Study data, including clinical and routine laboratory parameters, were collected and managed using REDCap electronic data capture tools hosted at Charité Universitätsmedizin Berlin (29, 30).

This study was approved by the Ethics Committee of Charité Universitätsmedizin Berlin (EA2/067/20, EA2/066/20) and is in accordance with the 1964 Declaration of Helsinki and its later amendments. The participants provided written informed consent.

### 2.2 Analysis of IgG response to EBV and human peptides by peptide CBA

#### 2.2.1 Cytometric bead array

Serum IgG binding to EBV EBNA epitopes and their respective human mimicry sequences was analyzed using a peptide cytometric bead assay (CBA). For this, BD functional CBA beads were first conjugated with streptavidin (SAV, Roche, Cat. no. 11721674001) using Sulfo-SMCC technology according to the manufacturer’s protocol (BD™ Cytometric Bead Array Functional Bead Conjugation Buffer Set, Cat. no. 558556). In brief, the disulfide bridges on functional beads were reduced with Dithiothreitol (Applichem, Cat. no. A1101,005), and in parallel, SAV was modified by reacting with Sulfo-SMCC (ThermoFisher Scientific, Cat. No. 22322). Following re-buffering using a Bio-Spin® 30 Tris Column (Bio-Rad, Cat. No. 7326231), the SAV-coated beads were stabilized by a final treatment with N-ethylmaleimide (ThermoFisher Scientific, Cat. no. 23030). After SAV binding was confirmed using a PE-coupled anti-SAV antibody (BioLegend®, Cat. no. 410504) by flow cytometry, the biotinylated 15-mer peptides were coupled to individual SAV-conjugated bead populations. Briefly, the SAV-conjugated beads were incubated with the corresponding biotinylated 15-mer peptides in a final concentration of 0.1 µM in Capture Bead Diluent (BD™ Cytometric Bead Array Human Soluble Protein Master Buffer Kit, Cat. no. 558265). Sequences of the EBV, the human mimicry, and the non-sense EBV EBNA1 scrambled (negative control) peptides are shown in Table II. Biotinylated peptides were synthesized via Fmoc-based solid phase peptide synthesis (SPPS) on polystyrene resin and purified by RP-HPLC by JPT Peptide Technologies GmbH (Berlin, Germany). A unique bead-labeling of APC and APC-Cy7 allowed multiplexing of up to 15 peptide-coupled bead populations.

**Table II.**
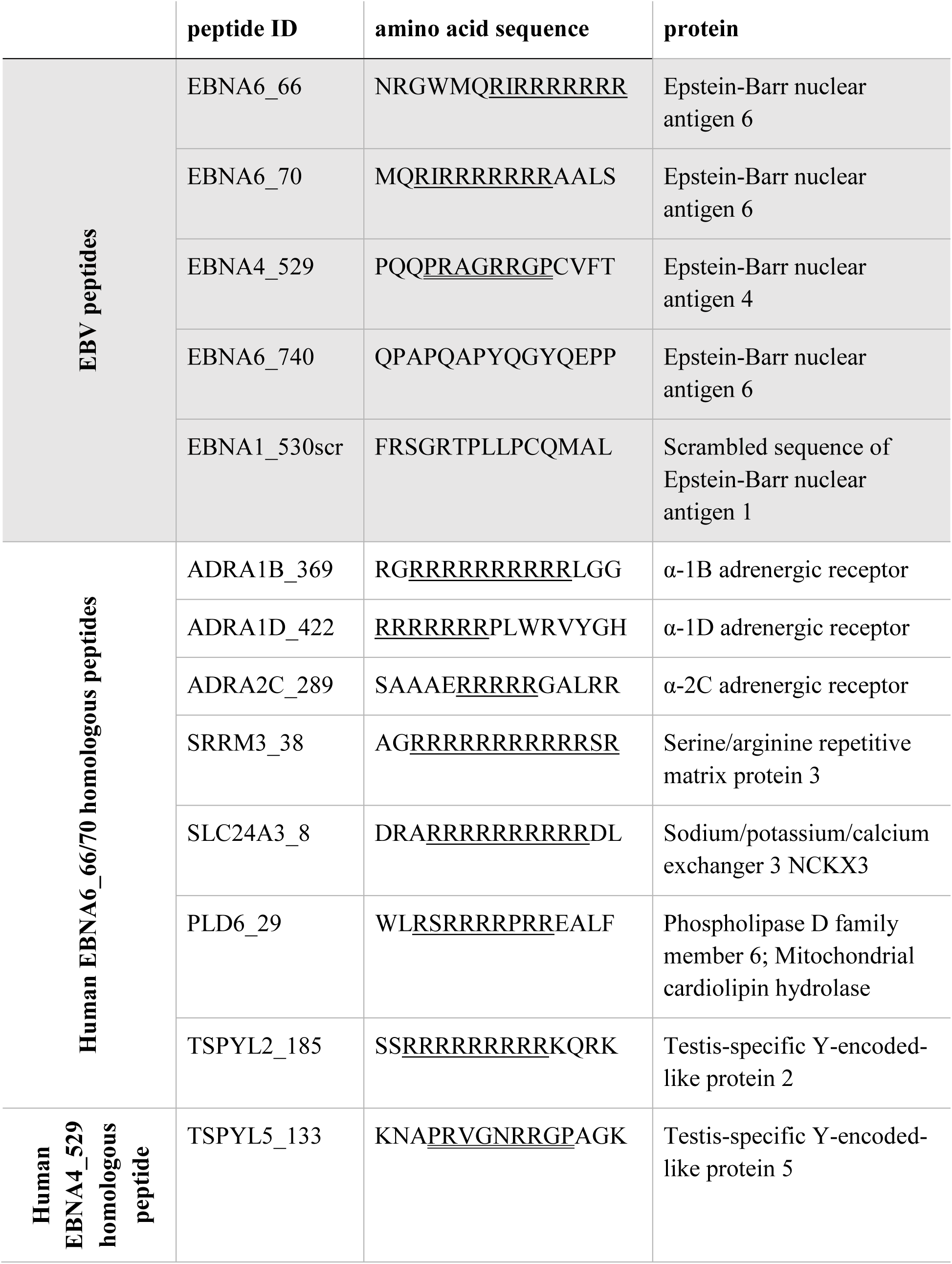
15-mer peptides of EBV and human sequences with homologous sequences underlined where the number after “_” in the peptide ID refers to the starting position of peptide in the reference protein sequence of EBV and human genome.

The measurement of patient and control serum IgG was performed according to the manufacturer’s protocol in a final serum dilution of 1:100 for one hour using Fcγ fragment specific R-Phycoerythrin AffiniPure™ Goat Anti-Human IgG (Jackson Immuno Research, Cat. no. 109-115-098) in a final dilution of 1:50 (BD™ Cytometric Bead Array Human Soluble Protein Master Buffer Kit, Cat. no. 558265). Samples were measured on a CytoFlex LX flow cytometer, data was acquired with CytExpert 2.4 and analyzed using FlowJo 10.8.2. Each bead set included a bead population coated with a scrambled sequence of EBV EBNA1 to control for unspecific IgG binding. The median fluorescence intensity (MFI) of detected unspecific binding was subtracted from each MFI of detected bound serum IgGs to peptide-coupled beads of interest. Since we found an intra-assay variability of up to 30 %, a positive response was defined as MFI > 30 % compared to the MFI signal of IgG binding to scrambled negative peptide (EBNA1_530scr) of the respective serum.

### 2.3 Analysis of IgG response to EBV and human full-length proteins

To detect serum IgG antibody responses to EBV proteins and potential human autoantigens, sera were tested in a multiplex dot blot assay as described (31). Briefly, 5 µl of concentration-adjusted recombinant His_6_-tagged proteins were spotted on a nitrocellulose membrane. Following a blocking step with 5% milk powder in PBS the membrane was co-incubated overnight at 4°C with an anti-His_6_ antibody (clone 3D5) and 1:1000 diluted serum in a 3% milk buffer. Following washing, membranes were incubated with fluorescence-labeled anti-mouse IgG antibody (LI-COR^®^ IRDye 680) and anti-human IgG antibody (LI-COR^®^ IRDye 800) to quantify the amount of spotted proteins and binding of human IgG. The membranes were scanned in a LI-COR^®^ Odyssey FC scanner that reports results as arbitrary fluorescence units (AFU), returning a CW700 and a CW800 reading for each dot on the membrane corresponding to the protein concentration (anti-His_6_) and the human serum response, respectively. A standard curve of recombinant His_6_-tagged human IgG, as well as solvent (8 M urea) and Ni-NTA agarose-affinity enriched mock-transfected HEK293T cell lysate, were used for specific standardization and background correction, enabling blot-to-blot comparability.

Autofluorescence signals caused by the nitrocellulose membrane or the solvent as well as any possible fluorescence due to serum responses against HEK293T proteins were subtracted from readings for antigenic proteins. Background-subtracted AFU values for proteins in the CW800 and CW700 channels were converted to normalized arbitrary values using a simple linear regression model drawn from values obtained for serial dilutions of recombinant IgG for each individual membrane. Next, the quotient of normalized CW800 and CW700 values was formed to compensate for potential differences in the amount of sample protein spotted on the membrane. This normalized AFU value describes sera IgG binding against EBV and human proteins. All normalized AFU values >0 were considered as positive IgG response.

### 2.4 Statistical analysis

Statistical data analyses were done using R Version 4.2.1 and GraphPad Prism Version 9.5.1. Patient characteristics are presented as median and range for continuous variables stratified by study group. Specific IgG binding was descriptively analyzed, presenting their medians and Interquartile Ranges (IQRs) for each study group. Univariate comparisons of two independent groups were done using the nonparametric Mann-Whitney test and of more independent groups using the Kruskal–Wallis test. Two-tailed tests were used with a significance level of 5%.

In addition, IgG binding was analyzed using linear models. To accommodate skewed distributions and outliers in the IgG levels, a log-transformation with a base of 2 was applied. Subsequently, linear models were fitted using the R-function ‘lmFit’ from the R-package ‘limma’, and model parameters were estimated utilizing the empirical Bayes method (R-function ‘eBayes’). The results were presented through volcano plots with a log2 fold change (FC) cutoff at one and a p-value cut-off of 0.1.

The frequency of positive IgG reactivity was expressed as a percentage. Pairwise comparisons of categorical variables between patient groups with HC group, respectively, was done using two-tailed Chi-Squared test with a significance level of 5%.

Correlations of IgG binding and clinical scores were analyzed by presenting heatmaps displaying Spearman-type correlation coefficients ≥ ±0.3 indicating at least small to moderate correlations (32). P-values less than 0.05 were considered as statistically significant and were indicated by * in the heatmaps.

## 3 Results

### 3.1 IgG reactivity against EBV and homologous human peptides

In a previous study using a regression model for binary outcomes, a significantly elevated IgG response was identified against two arginine-rich EBV EBNA peptides in piME/CFS patients compared to HC (23). Using the RefSeq_select and UniProtKB/Swiss-Prot databases, we identified proteins with high sequence homologies to EBNA6_70/66 and a putative role in ME/CFS: α-adrenergic receptors (ADRA1B, ADRA1D, ADRA2C), the sodium/potassium/calcium exchanger 3 (SLC24A3), the serine/arginine repetitive matrix protein 3 (SRRM3), the mitochondrial phospholipase D family member 6 (PLD6) and the testis-specific Y-encoded-like protein 2 (TSPYL2) (Table II). All these alignments share a characteristic poly-R sequence. For EBNA4_529, a homology was found with the testis-specific Y-encoded-like protein 5 (TSPYL5) containing three arginines (Table II).

In the present study, we analyzed antibody responses to these EBV EBNA and homologous human peptides, as well as to the corresponding full-length proteins in female PCS, pcME/CFS), piME/CFS patients and HC. Patients were selected for a similar age distribution Table I), but piME/CFS had a significantly longer disease duration (median 3.1 years) than pcME/CFS (median: 0.75 years) and PCS (median: 0.58 years) patients. As expected, ME/CFS patients suffered from a greater impairment and limitation of physical function, severe fatigue, and PEM compared to PCS patients. The EBNA6_740 peptide was included as a positive control because most patients and HCs had high antibody levels in our previous study (22). An overview of the study design is shown in Figure 1.

**Figure 1.**
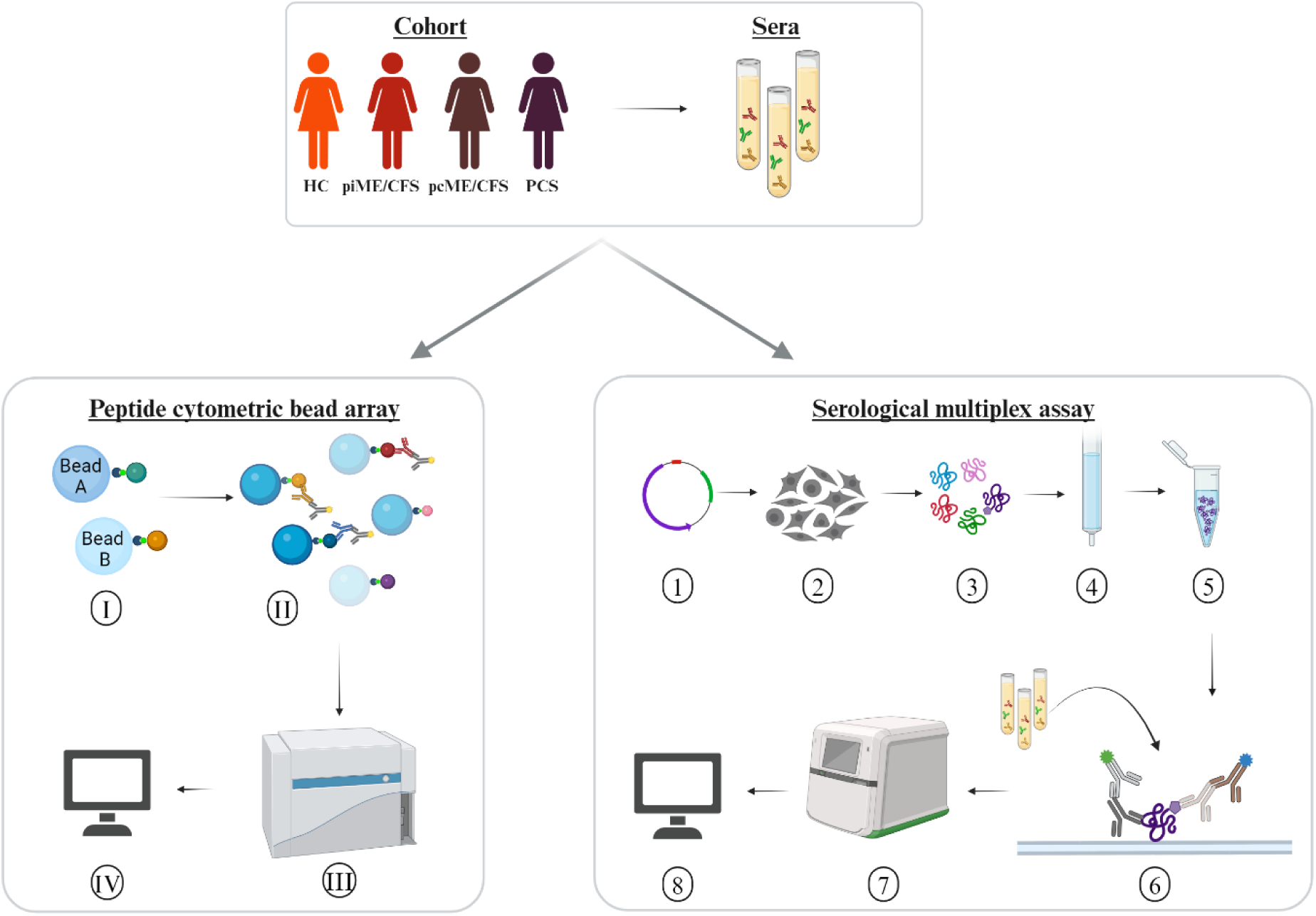
Experimental design. Sera of age-matched female patients with myalgic encephalomyelitis/chronic fatigue syndrome (ME/CFS) after SARS-CoV-2 (pcME/CFS), or after other infectious triggers (piME/CFS), as well as post-COVID syndrome (PCS) were analyzed in comparison to healthy controls (HC). Peptide cytometric bead array (CBA) for detection of IgG response to peptides of interest (I) Coupling of the beads with 15-mer peptides of interest. (II) Measurement of the serum IgG response against the peptides of interest in a multiplex approach. (III) Measurement of the serum IgG responses in the CytoFlex LX device. (IV) Data analysis of median fluorescence intensity (MFI). Serological multiplex array measurement of serum IgG responses to proteins of interest. (1) Expression vectors for C-terminally His6-tagged proteins of interest were generated using a pcDNA3.1-derived plasmid. (2) Transfection into HEK293T cells. (3) Harvesting and lysing cells after four days. (4) Purification of His6-tagged proteins over to Ni2+-NTA columns. (5) Verification of protein integrity and identity. (6) Addition of the proteins to a nitrocellulose membrane and incubation with an anti His6-tag monoclonal mouse antibody. (7) Detection of bound human and mouse antibodies with secondary antibodies in a near-infrared detection system. (8) Antibody responses to proteins were quantified by normalization of arbitrary fluorescence units (AFU) relative to the amount of spotted protein and a standard. Created in BioRender https://BioRender.com/p19y004

We observed IgGs reactive with EBNA6 and EBNA4 and with all the homologous human sequence peptides in sera of healthy controls and patients (Figure 2). A significantly increased IgG binding to the EBNA6_66/70 homologous sequences TSPYL2_185 and SRRM3_383 was found in PCS patients (Figure 2 A). Moreover, significantly higher levels of IgG antibodies reactive to the EBNA4_529 homolog TSPYL5_133 were found in all patient groups compared to HCs (Figure 2 B). Statistical analysis using linear models as implemented in Limma showed significantly elevated IgG reactivity to SRRM3_383, TSPYL2_185, and TSPYL5_133 in piME/CFS and PCS patients, and to TSPYL5_133 in the pcME/CFS cohort compared to HC (Figure 2 D-F).

**Figure 2.**
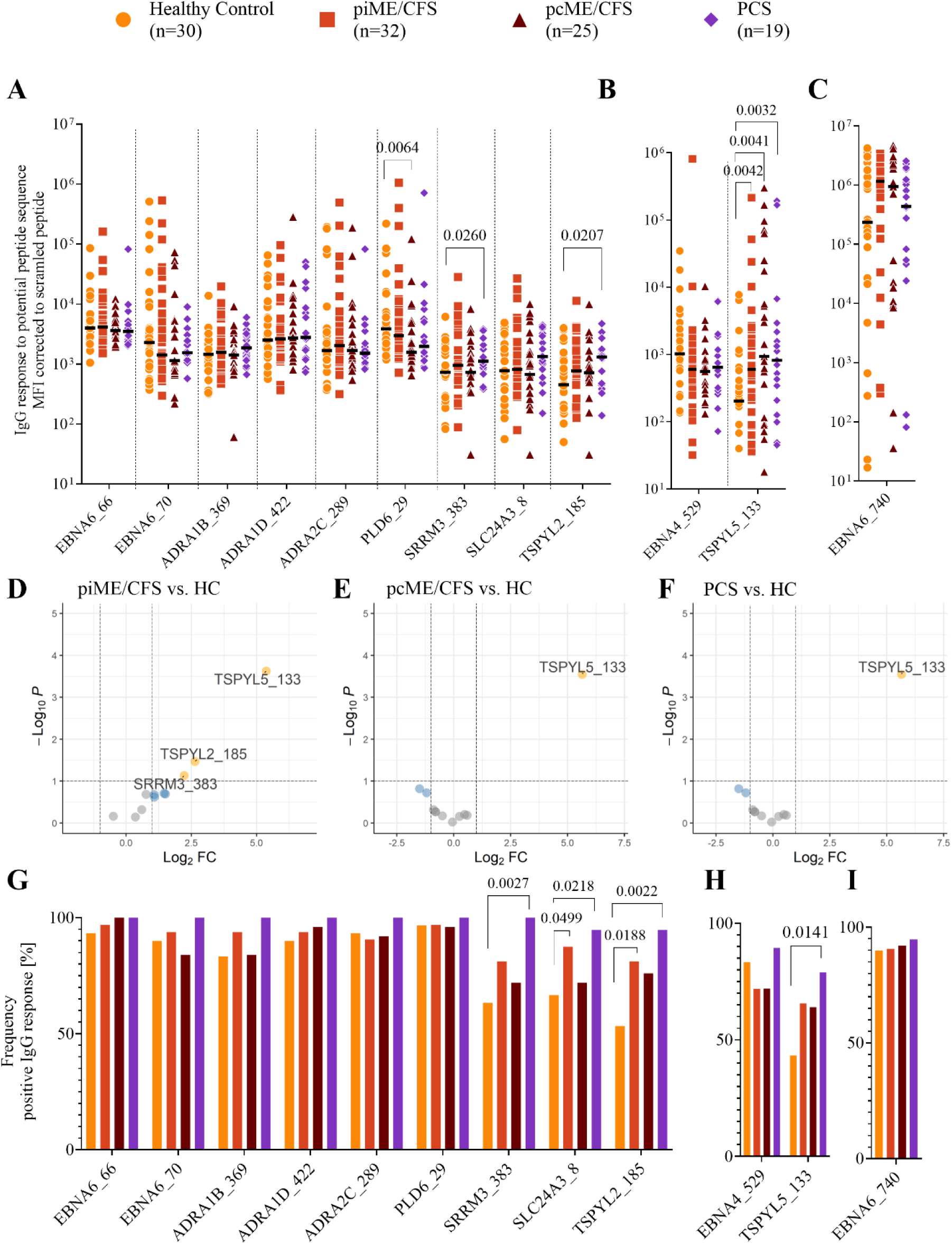
Analysis of IgG reactivity to peptides by CBA. Shown are the individual serum IgG responses as Mean Fluorescence Intensity (MFI) corrected to the MFI of scrambled peptide (A-F) and the frequency of positive IgG responses in % (G-H) of the whole patient cohorts (piME/CFS, pcME/CFS, PCS) and the healthy controls (HC). The IgG responses against (A, G) EBNA6_66/70 and homologous human peptides, (B, H) EBNA4_529 and homologous human peptide TSPYL5_133, as well as (C, I) EBNA6_740 were determined. Statistical comparison of the level of specific IgG binding between all patient cohorts and the HC cohort was done using the Mann-Whitney-U test (A-C). For the comparisons of IgG frequencies between study groups Chi-Squared test was used (G-I). P-values less than 0.05 of two-tailed tests were considered statistically significant. The Limma analysis (D-F) plotted for 12 variables and filtered for p-value <0.1. The enhanced volcano plots compare the different patient cohorts piME/CFS (D), pcME/CFS (E) and PCS (F), each to healthy controls (HC).

Looking at the percentage of subjects with specific IgG reactivity within the cohorts, we found antibodies to the EBNA6 poly-R sequences, as well as the ADRA and PLD6_23 peptides in almost all PCS patients, and most ME/CFS and HCs (Figure 2G). IgG reactivity to the SLC24A3_8, TSPYL2_185, and SRRM3_383 peptides was found in 40 – 70% of the HCs but again in 90 – 100% of PCS and significantly more frequent to TSPYL2_185 and SLC24A3_8 peptides in piME/CFS patients compared to HC, too (Figure 2G). IgG reactivity to EBNA4_529 was found at similar levels in about three-quarters of all samples, but more patients responded to the homologous peptide TSPYL5_133 (Figure 2 H). As in our previous study, a strong IgG reactivity against EBNA6_740 was found in 80 – 95% of all subjects (Figure 2 C, I).

We next investigated the potential cross-reactivity of IgG levels to EBV and homologous human polyR peptide sequences by Spearman‘s correlation coefficient (Figure 3 A-D). Positive correlations were observed for EBNA6_66/70 IgG with most human peptide IgGs in all cohorts. A correlation of EBNA4_529 and TSPYL5_133 was seen in pcME/CFS only. No correlations with the non-homologous EBN6_740 were seen.

**Figure 3.**
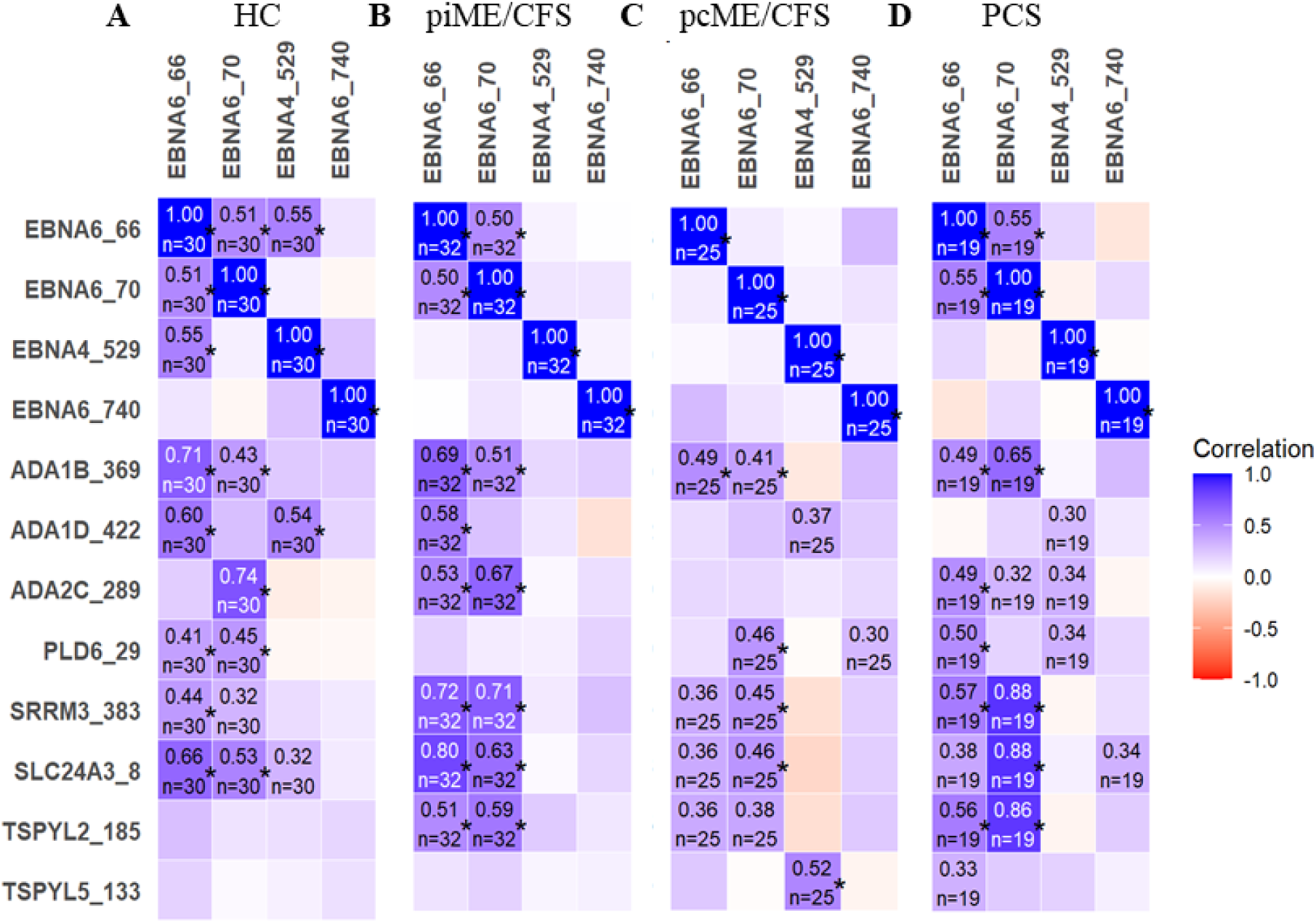
Heatmaps presenting the Spearman’s correlation coefficient r ≥ 0.3 of the correlation between the IgG reactivity to EBV EBNA peptides and homologous human peptides. (*p-values < 0.05)

### 3.2 IgG reactivity against EBNA and human full-length protein

Next, we studied the IgG responses to the EBV EBNA and human full-length proteins corresponding to the peptide sequences analyzed using a multiplex assay. As a specific cutoff for positive antibody responses towards these proteins has not been defined yet we show background corrected, normalized arbitrary fluorescence units (AFU) reactivity against each protein as described in the method section. We observed IgG binding to EBNA6 and EBNA4 proteins in most samples, which were higher against EBNA6 in patient cohorts compared to HC, with significance in pcME/CFS (Figure 4 A). Further IgG responses to the human proteins were seen in all study cohorts, although signals were low in many individuals. IgG responses to ADRA proteins were observed in 20 – 40% of the HCs but were significantly more frequent (Figure 4 C) and with higher titers in PCS and ME/CFS patients (Figure 4 A). The frequency of IgG reactive against TSPYL2 was similar in study groups (Figure 4 C), but both ME/CFS cohorts showed higher levels of autoantibodies to TSPYL2 compared to HCs (Figure 4 A). Similar levels and frequencies of antibodies binding to EBNA4 and TSPYL5 were observed in all study groups (Figure 4 B, D).

**Figure 4.**
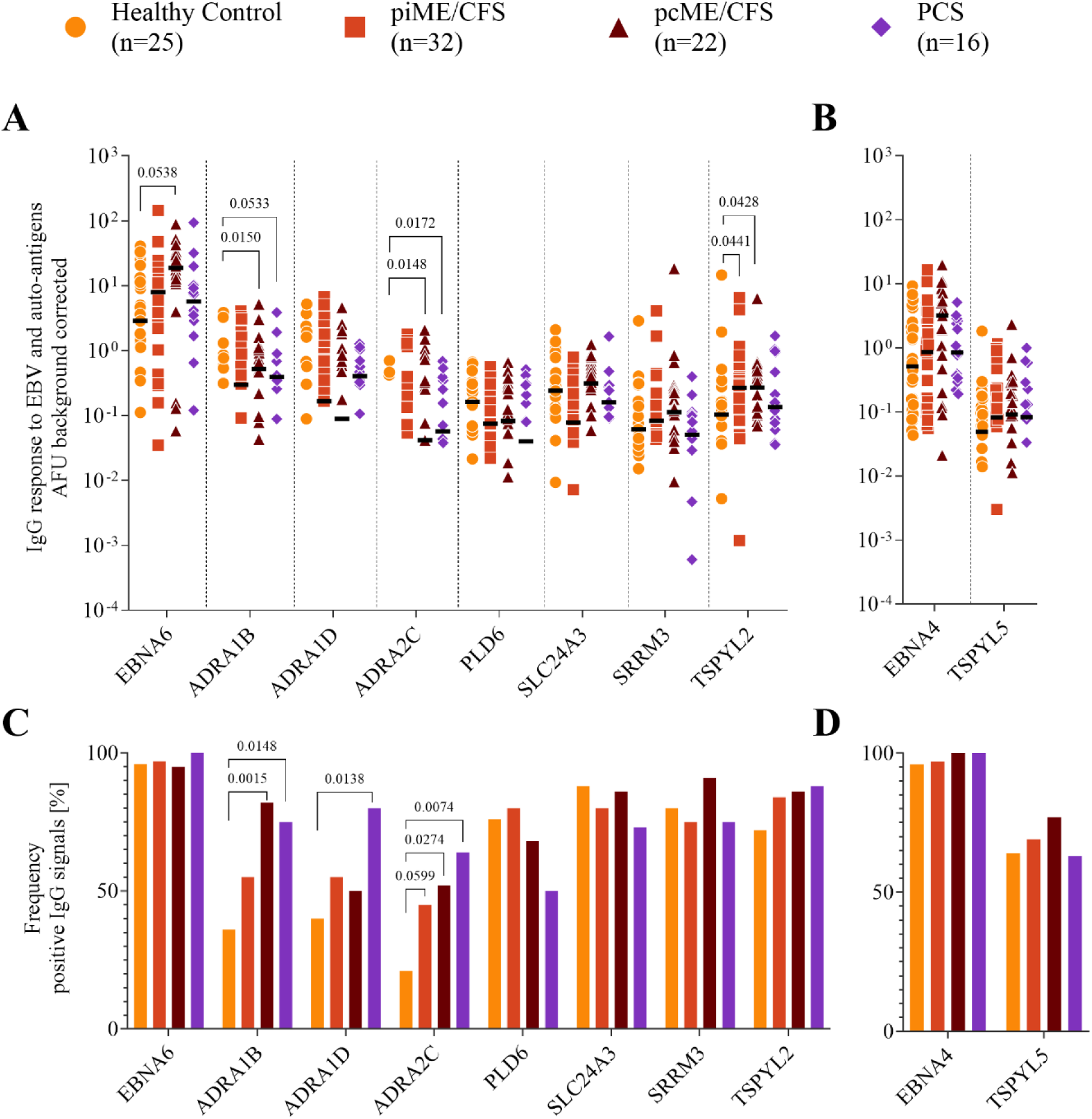
Analysis of IgG reactivity to proteins. Shown are (A-B) the individual serum IgG responses in arbitrary Unit (AFU) and (C-D) the frequency of positive IgG responses in % of the whole patient cohorts (piME/CFS, pcME/CFS, PCS) and the healthy controls (HC). The IgG responses against (A, C) EBNA6 and human proteins, (B, D) EBNA4 and human TSPYL5 proteins were determined. Statistical comparison of the level of specific IgG binding between patients and HC groups was done using the Mann-Whitney-U test (A-B). For the comparisons of IgG frequencies between study groups Chi-Squared test was used (C-D). A two-tailed p-value of <0.05 was considered statistically significant.

### 3.3 Association between IgG responses and symptom severity

Next, we investigated whether the levels of autoantibodies were associated with the severity of key symptoms in patients using Spearman correlation analyses (Figure 5). In PCS patients, we found significant positive correlations of most antibody levels, including those to EBNA6_66/70, with autonomic dysfunction assessed by COMPASS and of IgG to two ADRA peptides with fatigue and PEM. Several others correlated by trend (Figure 5 C). No significant correlations of antibodies to the respective proteins with symptoms were found in this patient group, but a trend of IgG to all three ADRA proteins with PEM severity is worth mentioning (Figure 5 F).

**Figure 5.**
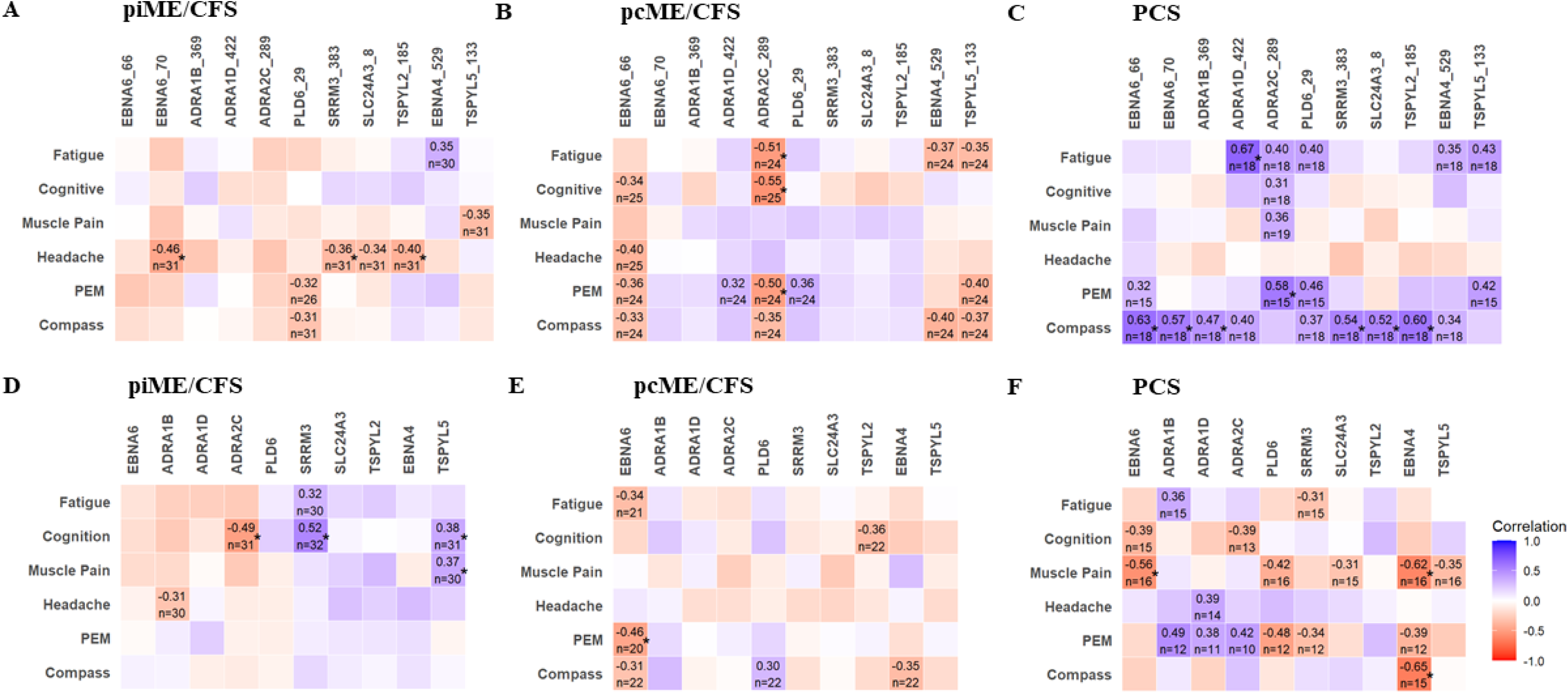
Associations of autoantibodies with symptom severity. IgG reactive to EBV and human peptides (A-C) and to corresponding proteins (D-E) were analyzed by Spearman correlation. Heatmaps presenting the Spearman correlation coefficient r ≥ 0.3 (*p-values < 0.05).

In the ME/CFS cohorts, no significant positive association between autoantibodies to peptides and symptom severity was observed (Figure 5 A,B). The piME/CFS group showed significant positive correlations of autoantibodies to SRRM3 protein with cognition and to TSPYL5 protein with both cognition and muscle pain (Figure 5 D). In contrast to PCS in pcME/CFS several inverse correlations between IgG responses to ADRA2C_422 peptides and symptoms, such as fatigue, cognition and PEM were found (Figure 5 B). Similar associations were also found when symptom severity was compared in patients with high versus low autoantibody levels (Supplemental Figure).

## 4 Discussion

This study characterized autoantibody responses to human peptides with high sequence homology to EBV arginine-rich (poly-R) sequences and to respective proteins in PCS and ME/CFS patients. Elevated levels of several autoantibodies in patients and positive correlations with the severity of key clinical symptoms point to their clinical relevance. The correlations of IgG responses to EBNA6_66/70 and human homologous poly-R peptides suggests cross-reactivity or molecular mimicry. In Figure 6 we propose a conceptual framework to explain the findings of our study. The associations of levels of several autoantibodies with symptom severity suggest that autoantibodies to poly-R sequences may affect the function of the corresponding proteins playing a role in autonomic, mitochondrial, and vascular function.

**Figure 6.**
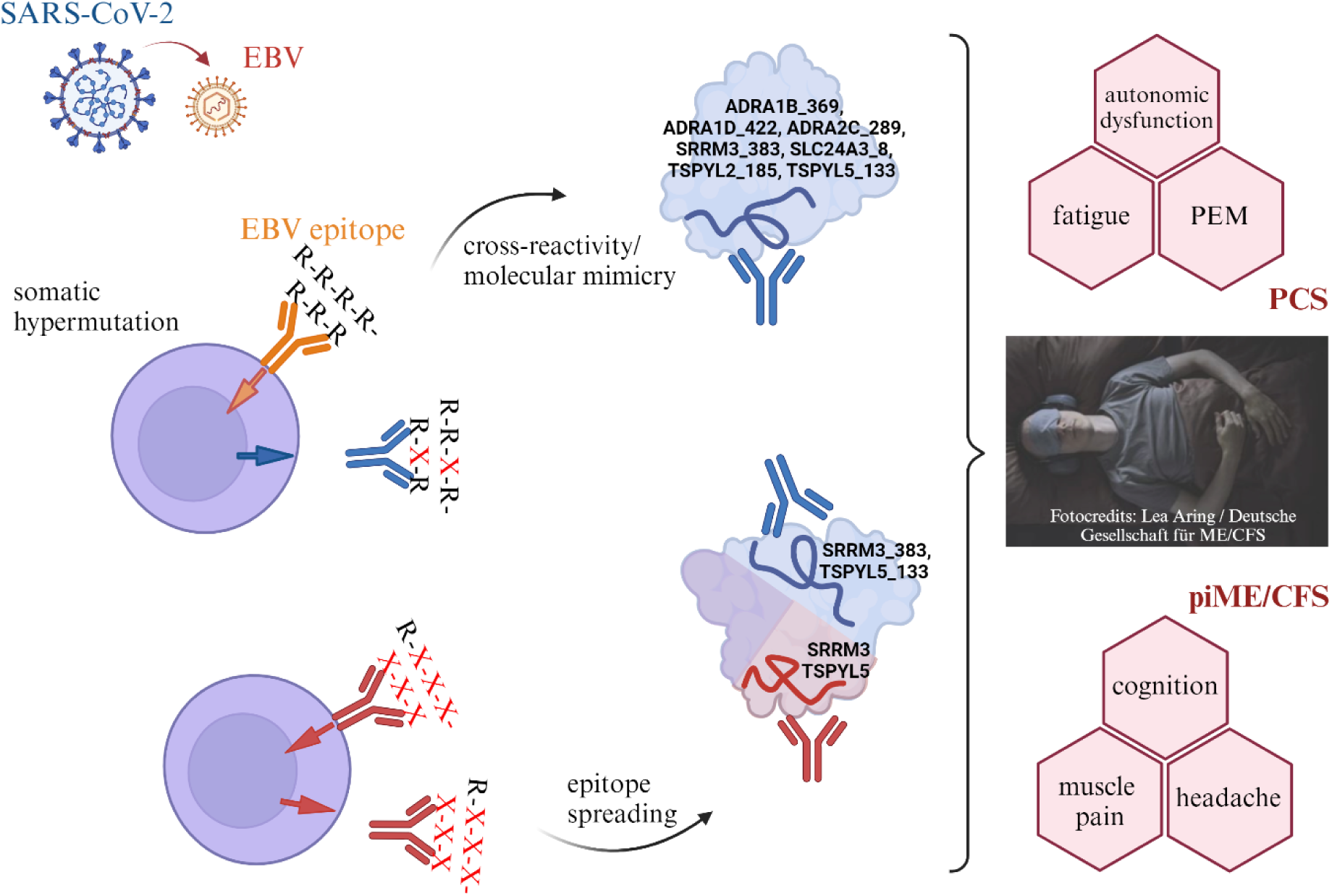
Proposal for a conceptual framework to explain the findings of this study. EBV reactivation which may be triggered by COVID-19 activates B cells, producing antibodies reactive to EBNA4 and EBNA6 arginine-rich sequences and corresponding human sequences. Somatic hypermutation can alter antibody specificities, which may induce or enhance cross-reactivity to poly-R sequences of human proteins. The autoantibody response to these poly-R epitopes and consecutive immune activation can stimulate B cells, producing autoantibodies to other epitopes of these self-antigens. Autoantibody binding may alter function of proteins and thus may lead to autonomic, mitochondrial or vascular dysfunction resulting in clinical symptoms. Created in BioRender. https://BioRender.com/m23y013

We observed clear differences in autoantibody titers and their association with symptoms among the three patient cohorts. In PCS highest levels and frequencies of autoantibodies to several human peptides were found. In line with this, autoantibodies to several poly-R peptides including ADRA correlated with the severity of autonomic dysfunction, fatigue, and PEM in PCS. More frequent autoantibodies to **ADRA** protein were found in all patient cohorts. Adrenergic receptors are important regulators of autonomic function, and a correlation of autoantibodies to adrenergic receptors with symptom severity was already shown for PCS and ME/CFS in several studies (7, 33). It is tempting to speculate that the inverse correlations between IgG responses to ADRA2C_422 and symptoms we found in pcME/CFS in contrast to PCS in this study may indicate epitope spreading (Figure 6).

Elevated levels of autoantibodies to **TSPYL2** peptide or protein were found in all patient cohorts. TSPYL2 is known to regulate TGF-β signaling and can inhibit Sirtuin-1 (SIRT1) activity which may result in mitochondrial dysfunction (34, 35). TGF-β levels have been reported to be elevated in ME/CFS patients in several studies (36). Higher autoantibodies to **SLC24A3** peptide were observed in PCS and piME/CFS, which is highly expressed in brain, skeletal, and smooth muscles and was shown to regulate cytoplasmic free Ca^2+^ and contractility in arterial smooth muscle (37). A single nucleotide variant in SLC24A3 was found to be a risk factor for migraine, a frequent comorbidity in PCS and ME/CFS (38).

Elevated autoantibodies to **SRRM3** and **TSPYL5** peptides were found in PCS and piME/CFS cohorts. In piME/CFS patients, we found autoantibodies against SRRM3 and TSPYL5 protein to be associated with cognitive impairment and for TSPYL5 in addition with muscle pain. Autoantibodies to these proteins may have been triggered initially via IgG reactive to the poly-R sequence peptides. Since we observed no correlation of IgG responses to corresponding peptides with symptoms in piME/CFS, epitope spreading may be a hypothetical mechanism (Figure 6). The correlations of autoantibodies and symptom severity are interesting when looking at the function of these proteins. SRRM3 expressed in the brain is a regulator of alternative splicing required for motor coordination and affects switching the gamma-aminobutyric acid (GABA) effect from excitatory to inhibitory (39, 40). Enhanced sensitivity to light and noise in ME/CFS is assumed to be due to low GABA levels. Noteworthy, a single nucleotide variant in SRRM3 was found to be associated with ME/CFS (41). In multiple sclerosis, autoantibodies to SRRM3 were found already years before the disease outbreak (42). TSPYL5 is known to be an angiogenic regulator playing a key role in maintaining endothelial integrity, function, and proliferation (43). Several studies showed hypoperfusion of the brain and muscle and disturbed microcirculation in ME/CFS (44–46). Recently, we have observed that serum from PCS patients, but not from ME/CFS patients, promotes endothelial tube formation *in vitro* (47). Thus, it is tempting to speculate that the observed association of TSPYL5 autoantibodies with cognitive impairment, headache, and muscle pain may be due to a potentially inhibitory effect on angiogenesis in piME/CFS. However, why such correlations could not be found in pcME/CFS remains unclear. The shorter disease duration of pcME/CFS compared to piME/CFS patients (median 0.75 versus 3.1 years) may be a possible explanation. Noteworthy, all self-antigens studied here are expressed in the brain, too. Autoantibodies against neuronal tissue were described in PCS in several studies (1, 9).

Poly-R motifs are present in many other viruses, including the Torque Teno virus (TTV), adenoviruses, and human papillomavirus HPV (48). Besides EBV, adenoviruses and HPV are known triggers of ME/CFS, too, and higher levels of TTV were recently described in ME/CFS (49). T cell clones reactive to the poly-R sequence in TTV were isolated from cerebrospinal fluid in a multiple sclerosis patient (48). These T cell clones also reacted to poly-R sequences of several human autoantigens, including ADRA1B/D and ADRA2C, but also dopamine D2 receptor, purinergic P2X2 receptor, inner mitochondrial membrane translocase TIM17, Toll-like receptor 9, and phosphatidylcholine transfer protein among others which may play a role in ME/CFS. These findings suggest that infection with or reactivation of other viruses than EBV containing arginine-rich motifs may also contribute to the autoantibody findings in our study. Therefore, we hypothesize that arginine-rich sequences occurring ubiquitously may result in frequent B and T cell activation and thus trigger autoreactivity through cross-reactivity, molecular mimicry, or epitope spreading (Figure 6).

## Limitations

Our study has several limitations. We included only females, and cohorts were rather small. The disease duration of the piME/CFS cohort was significantly longer than of the other two cohorts, which may have an influence on autoantibody findings. The correlation analyses of autoantibodies with symptom severity based on Spearman’s correlation coefficient is exploratory. Larger validation cohorts are also required for evaluating the diagnostic potential of these autoantibodies. We have no evidence that the IgGs reactive to the poly-R peptides cross-react with the native proteins which would require isolation of IgG to poly-R peptides and binding studies. Further functional studies with specific IgG autoantibodies in *in vitro* or animal models are required to proof their assumed functional role.

## Conclusion

Our findings provide evidence that IgG autoantibodies against peptides containing poly-R sequences and corresponding proteins are more frequent in PCS and ME/CFS. Further we observed several associations of autoantibody titers with severity of symptoms which may point to their functional relevance. Cross-reactivity with common viral motifs could explain the many different autoantibodies found in PCS and the broad clinical presentation. Our findings should prompt further studies into their pathogenic role, their potential use for diagnosis, and into drugs targeting autoantibodies.

## Abbreviations

ADRA: Alpha adrenergic receptor
AFU: Arbitrary fluorescence units
CBA: Cytometric bead array
EBNA: Epstein-Barr nuclear antigen
EBV: Epstein-Barr virus
GABA: Gamma-aminobutyric acid
GPCR: G protein-coupled receptors
HPV: Human papillomavirus
IgG: Immunoglobulin G
ME/CFS: Myalgic Encephalomyelitis/Chronic Fatigue Syndrome
MFI: Median fluorescence intensity
MRI: Magnetic resonance imaging
Pc: Post-COVID
PCS: Post-COVID syndrome
PEM: Post-exertional malaise
pi: Postinfectious
Poly-R: Poly arginine
SARS-CoV-2: Severe acute respiratory syndrome coronavirus 2
SF-36: Short form 36
SIRT1: Sirtuin-1
SLC24A3: Solute carrier family 24 member 3
Sulfo-SMCC: Sulfosuccinimidyl 4-(N-maleimidomethyl) cyclohexane-1-carboxylate
SRRM3: Serine/arginine repetitive matrix 3
TSPYL2/5: Testis-specific Y-encoded-like protein 2/-5
TTV: Torque-Teno-virus

## Data Availability

All data produced in the present study are available upon reasonable request to the authors

## Others

We would like to thank the patients who participated in this study, as well as Anja Hagemann, Silvia Thiel, and Beate Follendorf for their work in patient care and data management.

## Authorship

FH, KF and SB conducted the laboratory work. FH, KF, KR, FK and FS analyzed the data of the study. CK, ES, KW, JBS, and CS diagnosed and enrolled the patients. FS, HF, and CH managed clinical and routine laboratory data. CS, FS, and JM provided concept, design, resources, and supervision. CS, FS, FH, MS, JM, UB, and NS interpreted and discussed the results. CS, FS, FH, NS, MS, JM and UB provided scientific insights. FH, CS, and FS wrote the manuscript. All authors have read, revised, and approved the final version of the manuscript.

## Submission declaration

This work has not been previously published and is not being considered for publication elsewhere. If accepted, this work will not be published elsewhere in the same form, in English, or any other language, including electronically without the written consent of the copyright holder.

## Declaration of Generative AI and AI-assisted technologies in the writing process

During the preparation of this work, the authors did not use generative AI and AI-assisted technologies in the writing process, which go beyond improving readability and language.

## References

1. Franke C, Boesl F, Goereci Y, Gerhard A, Schweitzer F, Schroeder M, et al. Association of cerebrospinal fluid brain-binding autoantibodies with cognitive impairment in post-COVID-19 syndrome. Brain, behavior, and immunity. 2023;109:139–43.

2. Woodruff MC, Ramonell RP, Haddad NS, Anam FA, Rudolph ME, Walker TA, et al. Dysregulated naive B cells and de novo autoreactivity in severe COVID-19. Nature. 2022;611(7934):139-47.

3. Jernbom AF, Skoglund L, Pin E, Sjöberg R, Tegel H, Hober S, et al. Prevalent and persistent new-onset autoantibodies in mild to severe COVID-19. Nat Commun. 2024;15(1):8941.

4. Boesl F, Goereci Y, Gerhard A, Bremer B, Raeder V, Schweitzer F, et al. Cerebrospinal fluid findings in patients with neurological manifestations in post-COVID-19 syndrome. Journal of Neurology. 2024;271(1):59–70.

5. Bodansky A, Wang C-Y, Saxena A, Mitchell A, Takahashi S, Anglin K, et al. Autoantigen profiling reveals a shared post-COVID signature in fully recovered and Long COVID patients. medRxiv. 2023:2023.02.06.23285532.

6. Sotzny F, Filgueiras IS, Kedor C, Freitag H, Wittke K, Bauer S, et al. Dysregulated autoantibodies targeting vaso-and immunoregulatory receptors in Post COVID Syndrome correlate with symptom severity. Front Immunol. 2022;13:981532.

7. Seibert FS, Stervbo U, Wiemers L, Skrzypczyk S, Hogeweg M, Bertram S, et al. Severity of neurological Long-COVID symptoms correlates with increased level of autoantibodies targeting vasoregulatory and autonomic nervous system receptors. Autoimmun Rev. 2023;22(11):103445.

8. Chen H-J, Appelman B, Willemen H, Bos A, Prado J, Geyer CE, et al. Transfer of IgG from Long COVID patients induces symptomology in mice. bioRxiv. 2024:2024.05.30.596590.

9. Santos Guedes de Sa K, Silva J, Bayarri-Olmos R, Brinda R, Alec Rath Constable R, Colom Diaz PA, et al. A causal link between autoantibodies and neurological symptoms in long COVID. medRxiv. 2024.

10. Kedor C, Freitag H, Meyer-Arndt L, Wittke K, Hanitsch LG, Zoller T, et al. A prospective observational study of post-COVID-19 chronic fatigue syndrome following the first pandemic wave in Germany and biomarkers associated with symptom severity. Nature Communications. 2022;13(1):5104.

11. Bonilla H, Quach TC, Tiwari A, Bonilla AE, Miglis M, Yang PC, et al. Myalgic Encephalomyelitis/Chronic Fatigue Syndrome is common in post-acute sequelae of SARS-CoV-2 infection (PASC): Results from a post-COVID-19 multidisciplinary clinic. Front Neurol. 2023;14:1090747.

12. Sotzny F, Blanco J, Capelli E, Castro-Marrero J, Steiner S, Murovska M, et al. Myalgic Encephalomyelitis/Chronic Fatigue Syndrome - Evidence for an autoimmune disease. Autoimmunity reviews. 2018;17(6):601–9.

13. Ryabkova VA, Gavrilova NY, Poletaeva AA, Pukhalenko AI, Koshkina IA, Churilov LP, et al. Autoantibody Correlation Signatures in Fibromyalgia and Myalgic Encephalomyelitis/Chronic Fatigue Syndrome: Association with Symptom Severity. Biomedicines. 2023;11(2).

14. Loebel M, Grabowski P, Heidecke H, Bauer S, Hanitsch LG, Wittke K, et al. Antibodies to beta adrenergic and muscarinic cholinergic receptors in patients with Chronic Fatigue Syndrome. Brain, behavior, and immunity. 2016;52:32–9.

15. Freitag H, Szklarski M, Lorenz S, Sotzny F, Bauer S, Philippe A, et al. Autoantibodies to Vasoregulative G-Protein-Coupled Receptors Correlate with Symptom Severity, Autonomic Dysfunction and Disability in Myalgic Encephalomyelitis/Chronic Fatigue Syndrome. Journal of Clinical Medicine. 2021;10(16):3675.

16. Kimura Y, Sato W, Maikusa N, Ota M, Shigemoto Y, Chiba E, et al. Free-water-corrected diffusion and adrenergic/muscarinic antibodies in myalgic encephalomyelitis/chronic fatigue syndrome. Journal of Neuroimaging. 2023;33(5):845–51.

17. Gravelsina S, Vilmane A, Svirskis S, Rasa-Dzelzkaleja S, Nora-Krukle Z, Vecvagare K, et al. Biomarkers in the diagnostic algorithm of myalgic encephalomyelitis/chronic fatigue syndrome. Front Immunol. 2022;13:928945.

18. Ruiz-Pablos M, Paiva B, Montero-Mateo R, Garcia N, Zabaleta A. Epstein-Barr Virus and the Origin of Myalgic Encephalomyelitis or Chronic Fatigue Syndrome. Front Immunol. 2021;12:656797.

19. Peluso MJ, Deveau TM, Munter SE, Ryder D, Buck A, Beck-Engeser G, et al. Chronic viral coinfections differentially affect the likelihood of developing long COVID. J Clin Invest. 2023;133(3).

20. Vojdani A, Vojdani E, Saidara E, Maes M. Persistent SARS-CoV-2 Infection, EBV, HHV-6 and Other Factors May Contribute to Inflammation and Autoimmunity in Long COVID. Viruses. 2023;15(2).

21. Jensen MA, Dafoe ML, Wilhelmy J, Cervantes L, Okumu AN, Kipp L, et al. Catalytic Antibodies May Contribute to Demyelination in Myalgic Encephalomyelitis/Chronic Fatigue Syndrome. Biochemistry. 2024;63(1):9–18.

22. Loebel M, Eckey M, Sotzny F, Hahn E, Bauer S, Grabowski P, et al. Serological profiling of the EBV immune response in Chronic Fatigue Syndrome using a peptide microarray. PLoS One. 2017;12(6):e0179124.

23. Sepulveda N, Malato J, Sotzny F, Grabowska AD, Fonseca A, Cordeiro C, et al. Revisiting IgG Antibody Reactivity to Epstein-Barr Virus in Myalgic Encephalomyelitis/Chronic Fatigue Syndrome and Its Potential Application to Disease Diagnosis. Front Med (Lausanne). 2022;9:921101.

24. Fonseca A, Szysz M, Ly HT, Cordeiro C, Sepúlveda N. IgG Antibody Responses to Epstein-Barr Virus in Myalgic Encephalomyelitis/Chronic Fatigue Syndrome: Their Effective Potential for Disease Diagnosis and Pathological Antigenic Mimicry. Medicina (Kaunas). 2024;60(1).

25. Bell DS. The doctor’s guide to chronic fatigue syndrome: understanding, treating, and living with CFIDS. (No Title). 1995.

26. Ware JE, Jr., Sherbourne CD. The MOS 36-item short-form health survey (SF-36). I. Conceptual framework and item selection. Med Care. 1992;30(6):473–83.

27. Cotler J, Holtzman C, Dudun C, Jason LA. A Brief Questionnaire to Assess Post-Exertional Malaise. Diagnostics (Basel). 2018;8(3).

28. Sletten DM, Suarez GA, Low PA, Mandrekar J, Singer W. COMPASS 31: a refined and abbreviated Composite Autonomic Symptom Score. Mayo Clin Proc. 2012;87(12):1196–201.

29. Harris PA, Taylor R, Thielke R, Payne J, Gonzalez N, Conde JG. Research electronic data capture (REDCap)--a metadata-driven methodology and workflow process for providing translational research informatics support. J Biomed Inform. 2009;42(2):377–81.

30. Harris PA, Taylor R, Minor BL, Elliott V, Fernandez M, O’Neal L, et al. The REDCap consortium: Building an international community of software platform partners. J Biomed Inform. 2019;95:103208.

31. Nuckel J, Planatscher E, Mohr AW, Deichl K, Mijocevic H, Feuerherd M, et al. Association between IgG responses against the nucleocapsid proteins of alphacoronaviruses and COVID-19 severity. Front Immunol. 2022;13:889836.

32. Cohen J. Statistical Power Analysis for the Behavioral Sciences. 2nd ed. ed. New York: Routledge; 1988 1 July 1988.

33. Ceccarini MR, Bonetti G, Medori MC, Dhuli K, Tezzele S, Micheletti C, et al. Autoantibodies in patients with post-COVID syndrome: a possible link with severity? Eur Rev Med Pharmacol Sci. 2023;27(6 Suppl):48-56.

34. Toh B-H, Tu Y, Cao Z, Cooper ME, Chai Z. Role of Cell Division Autoantigen 1 (CDA1) in Cell Proliferation and Fibrosis. Genes. 2010;1(3):335–48.

35. Pham Y, Tu Y, Wu T, Allen TJ, Calkin AC, Watson AM, et al. Cell division autoantigen 1 plays a profibrotic role by modulating downstream signalling of TGF-β in a murine diabetic model of atherosclerosis. Diabetologia. 2010;53(1):170–9.

36. Blundell S, Ray KK, Buckland M, White PD. Chronic fatigue syndrome and circulating cytokines: A systematic review. Brain, behavior, and immunity. 2015;50:186–95.

37. Dong H, Jiang Y, Triggle CR, Li X, Lytton J. Novel role for K+-dependent Na+/Ca2+ exchangers in regulation of cytoplasmic free Ca2+ and contractility in arterial smooth muscle. Am J Physiol Heart Circ Physiol. 2006;291(3):H1226–35.

38. Pisanu C, Preisig M, Castelao E, Glaus J, Pistis G, Squassina A, et al. A genetic risk score is differentially associated with migraine with and without aura. Human Genetics. 2017;136(8):999–1008.

39. Ciampi L, Mantica F, López-Blanch L, Permanyer J, Rodriguez-Marín C, Zang J, et al. Specialization of the photoreceptor transcriptome by ˂I˃Srrm3˂/I˃-dependent microexons is required for outer segment maintenance and vision. Proceedings of the National Academy of Sciences. 2022;119(29):e2117090119.

40. Nakano Y, Wiechert S, Bánfi B. Overlapping Activities of Two Neuronal Splicing Factors Switch the GABA Effect from Excitatory to Inhibitory by Regulating REST. Cell Reports. 2019;27(3):860–71.e8.

41. Schlauch KA, Khaiboullina SF, De Meirleir KL, Rawat S, Petereit J, Rizvanov AA, et al. Genome-wide association analysis identifies genetic variations in subjects with myalgic encephalomyelitis/chronic fatigue syndrome. Translational Psychiatry. 2016;6(2):e730-e.

42. Zamecnik CR, Sowa GM, Abdelhak A, Dandekar R, Bair RD, Wade KJ, et al. An autoantibody signature predictive for multiple sclerosis. Nature Medicine. 2024;30(5):1300–8.

43. Na H-J, Yeum CE, Kim H-S, Lee J, Kim JY, Cho YS. TSPYL5-mediated inhibition of p53 promotes human endothelial cell function. Angiogenesis. 2019;22(2):281–93.

44. Haffke M, Freitag H, Rudolf G, Seifert M, Doehner W, Scherbakov N, et al. Endothelial dysfunction and altered endothelial biomarkers in patients with post-COVID-19 syndrome and chronic fatigue syndrome (ME/CFS). Journal of translational medicine. 2022;20(1):138.

45. Sandvik MK, Sorland K, Leirgul E, Rekeland IG, Stavland CS, Mella O, et al. Endothelial dysfunction in ME/CFS patients. PLoS One. 2023;18(2):e0280942.

46. Charfeddine S, Ibn Hadj Amor H, Jdidi J, Torjmen S, Kraiem S, Hammami R, et al. Long COVID 19 Syndrome: Is It Related to Microcirculation and Endothelial Dysfunction? Insights From TUN-EndCOV Study. Front Cardiovasc Med. 2021;8:745758.

47. Flaskamp L, Roubal C, Uddin S, Sotzny F, Kedor C, Bauer S, et al. Serum of Post-COVID-19 Syndrome Patients with or without ME/CFS Differentially Affects Endothelial Cell Function In Vitro. Cells. 2022;11(15):2376.

48. Sospedra M, Zhao Y, zur Hausen H, Muraro PA, Hamashin C, de Villiers EM, et al. Recognition of conserved amino acid motifs of common viruses and its role in autoimmunity. PLoS Pathog. 2005;1(4):e41.

49. Giménez-Orenga K, Martín-Martínez E, Oltra E. Over-Representation of Torque Teno Mini Virus 9 in a Subgroup of Patients with Myalgic Encephalomyelitis/Chronic Fatigue Syndrome: A Pilot Study. Pathogens. 2024;13(9).

