## Supplemental data for "Autoantibodies to Arginine-rich Sequences Mimicking Epstein-Barr Virus in Post-COVID and Myalgic Encephalomyelitis/Chronic Fatigue Syndrome"

### Methods

*Statistical analysis*

In addition to correlation analysis, we conducted Mann-Whitney test to assess differences in the clinical parameters between two groups. The specific IgG responses were categorized into two groups: "High" and "Low", based on the median value of the data. If the median was found to be 0, all values equal to or less than 0 were classified as "Low", while values greater than 0 were categorized as "High".

### Results

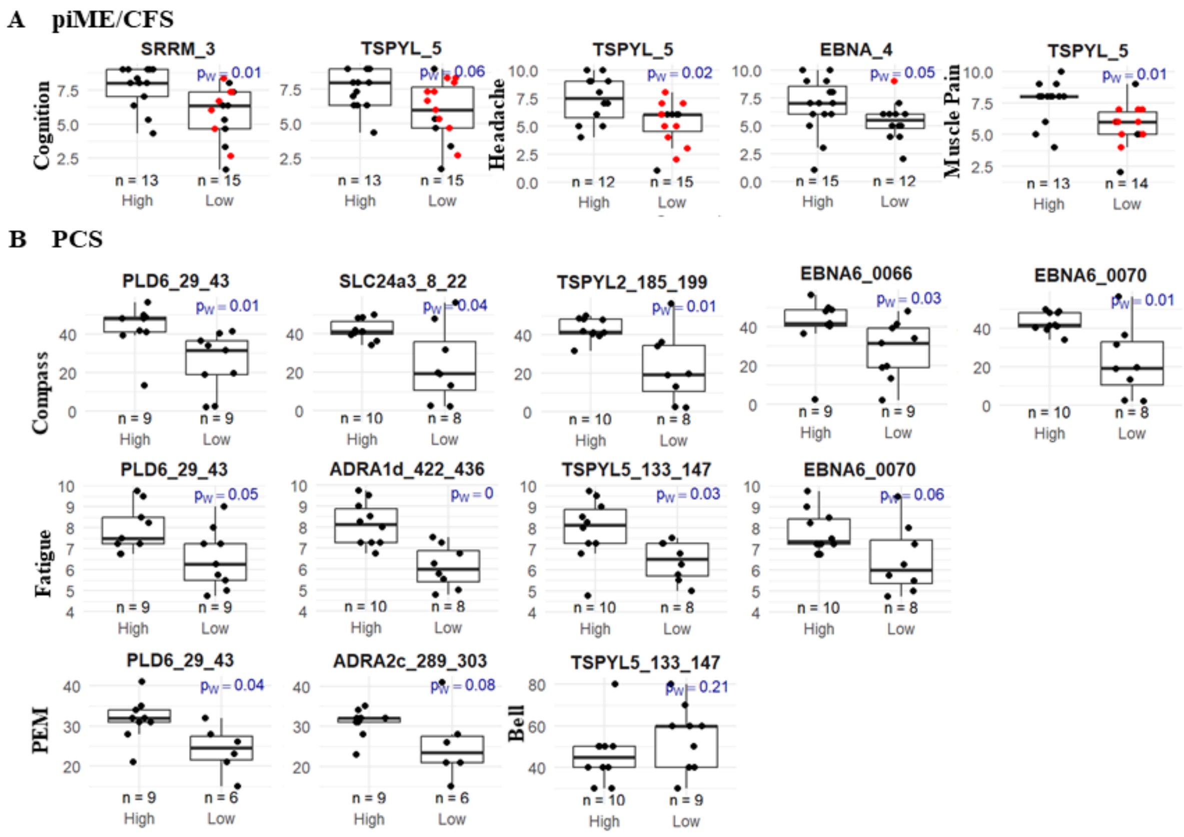


**Supplementary Figure: Associations of autoantibodies with symptom severity.** Shown are the clinical scores of patients with piME/CFS **(A)** or PCS **(A)** as median with interquartile ranges (IQR) comparatively for patient group with high or low IgG responses to human peptides or to corresponding proteins. Statistical comparison was done using the Mann-Whitney-U test. A two-tailed p-value of <0.05 was considered statistically significant. Samples without specific IgG binding were indicated as red dots.
